# Exposure to Noise and Cardiovascular Disease in a Nationwide US Prospective Cohort Study of Women

**DOI:** 10.1101/2023.06.07.23291083

**Authors:** Charlotte Roscoe, Stephanie T. Grady, Jaime E. Hart, Hari S. Iyer, JoAnn E. Manson, Kathryn M. Rexrode, Eric B. Rimm, Francine Laden, Peter James

## Abstract

**Background:** Long-term noise exposure is associated with cardiovascular disease (CVD), including acute cardiovascular events such as myocardial infarction and stroke. However, longitudinal cohort studies of long-term noise and CVD are almost exclusively from Europe, and few modelled nighttime and daytime noise separately. We aimed to examine the prospective association of outdoor long-term nighttime and daytime noise from anthropogenic sources with incident CVD using a US-based, nationwide cohort of women.

**Methods:** We linked L50 (median) nighttime and L50 daytime modelled anthropogenic noise estimates from a US National Park Service model to geocoded residential addresses of 114,116 participants in the Nurses’ Health Study. We used time-varying Cox proportional hazards models to estimate risk of incident CVD, coronary heart disease (CHD), and stroke associated with long-term average noise exposure, adjusted for potential individual- and area-level confounders and CVD risk factors (1988-2018). We assessed effect modification by population density, region, air pollution, vegetation cover, and neighborhood socioeconomic status, and explored mediation by self-reported average nightly sleep duration.

**Results:** Over 2,544,035 person-years, there were 10,331 incident CVD events. In fully-adjusted models, the hazard ratios for each interquartile range increase in L50 nighttime noise (3.67 dBA) and L50 daytime noise (4.35 dBA), respectively, were 1.04 (95% CI 1.02, 1.06) and 1.04 (95% CI 1.02, 1.07). Similar associations were observed for CHD and stroke. Stratified analyses suggested that associations of nighttime and daytime noise with CVD did not differ by prespecified effect modifiers. We found no evidence that inadequate sleep (< 5 hours per night) mediated associations of noise and CVD.

**Discussion:** Outdoor median nighttime and daytime noise at the residential address was associated with a small increase in CVD risk in a cohort of adult female nurses.

## Introduction

Noise, or unwanted sound exposure, is the second largest environmental cause of health problems, after air pollution, and has been associated with multiple adverse outcomes, which include annoyance, sleep disturbance, and poor concentration.^1–3^ In addition, epidemiological studies have found associations of long-term noise exposure with risk of metabolic and cardiovascular disease (CVD).^4^

Exposure to noise has been linked to short-term changes in circulation, such as blood pressure, heart rate, cardiac output, and vasoconstriction.^5–7^ These biological changes can occur not only at high sound levels in occupational settings, but also during exposure to lower levels of environmental noise in residential settings.^8^ Noise exposure activates the central nervous system and triggers a host of changes in various subsystems in the human body identical to a typical stress response.^9^ Reactions include activation of the hypothalamic-pituitary-adrenal axis and the sympathetic nervous system, which is triggered by limbic activity in the brain, and results in the release of stress hormones (glucocorticoids and catecholamines); repeated release of stress hormones associated with long-term noise exposure may manifest in inflammation, atherosclerosis, insulin-resistance, and CVD.^10–12^

Prospective cohort studies conducted in Europe^13–18^ and Canada^19^ suggest that higher levels of environmental noise are linked to increased risk of CHD and stroke, although findings are inconsistent.^20–23^ In a 2018 WHO review, evidence linking long-term noise exposure with CHD and stroke was graded as high- and moderate-quality, respectively^24^; however, few studies have been conducted in the US, and no study has assessed the prospective association of nighttime anthropogenic noise and CVD in a nationwide US study. Epidemiologic studies indicate that nighttime noise exposure may be more relevant for cardiovascular outcomes than daytime exposure.^25^ While many epidemiological studies have weighted noise models to assign a penalty to nighttime noise (e.g., adding a 10 dB to noise between 10pm and 7am), few studies specifically assessed nighttime noise exposure, which occurs when individuals are most likely to be home and sleeping.

In the US, anthropogenic noise has been assessed ecologically,^26^ however, there is a deficit of individual-level epidemiological US-based studies on noise and chronic disease. Using data from the nationwide, US-based Nurses’ Health Study (NHS) from 1988-2018, we examined the association of anthropogenic nighttime and daytime noise with CVD incidence. We also examined whether the association of noise and CVD differed by population density, region, air pollution level, greenness level, or neighborhood socioeconomic status (SES), and if associations were mediated by sleep.

## Methods

### Population

The Nurses’ Health Study (NHS) is a prospective cohort study designed to assess risk factors for chronic disease among women. In 1976, 121,701 female registered nurses aged 30-55 years, from 11 states (CA, CT, IN, IA, KY, MA, MI, MO, NY, NC, OH, PA, SC, and TX), returned an initial health-related questionnaire and have since been followed with biennial questionnaires on demographic and physical characteristics, lifestyle, and health status. Response rates at each questionnaire cycle have consistently been ≥90%.^27, 28^ The residential addresses have been geocoded (i.e., assigned latitude and longitude) and updated at each move of address. NHS participants currently reside in all states in the contiguous US and the District of Columbia (**Figure 1**). This analysis was conducted among all women who were alive and had no prior CVD in 1988 and had at least one residential address geocoded in the contiguous US for exposure assessment between 1988 and 2018. This study was approved by the Institutional Review Board of Brigham and Women’s Hospital, Boston, MA.

**Figure 1.**
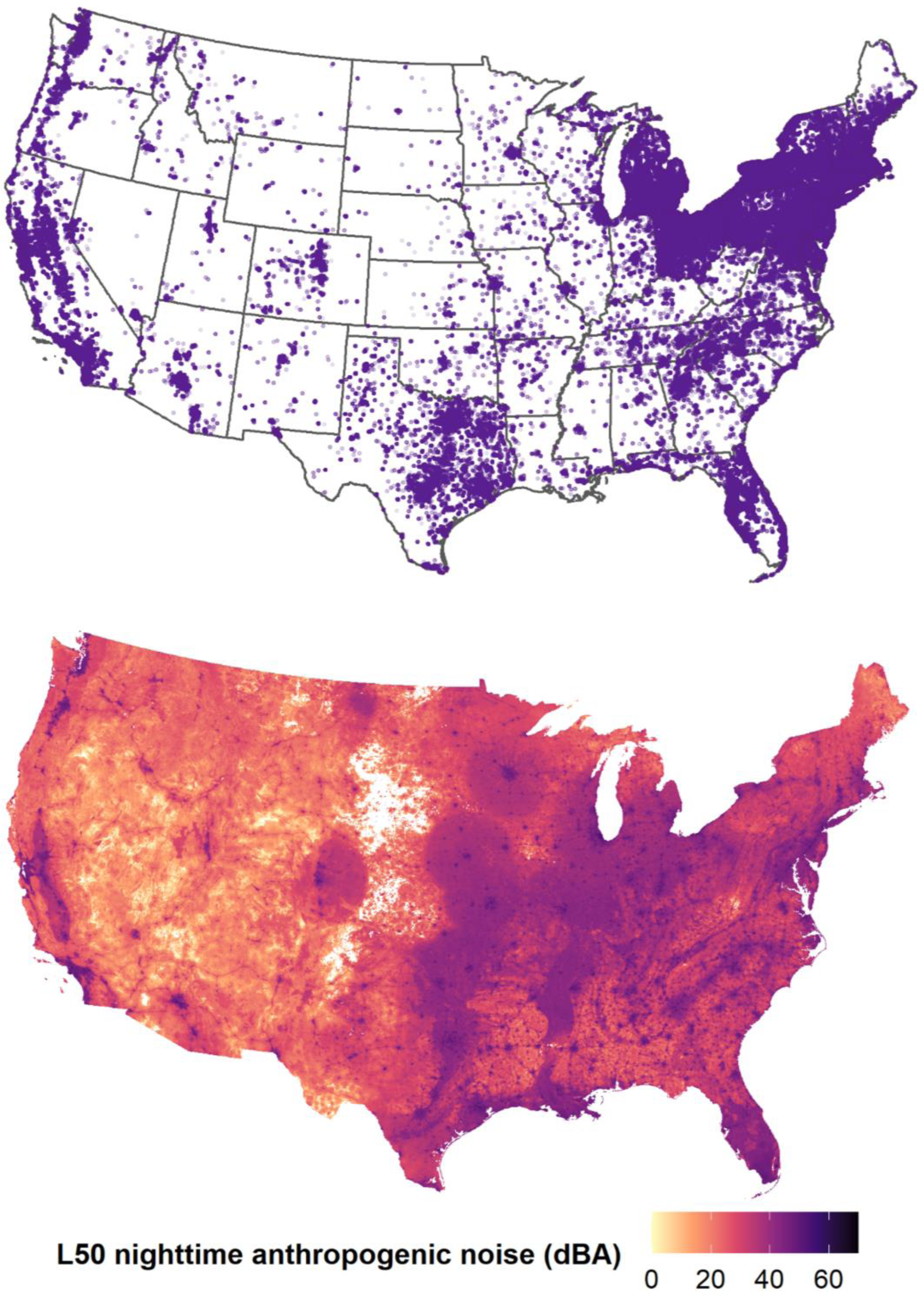
Location of NHS Addresses from 1988-2018 (above) and Map of Anthropogenic Nighttime L50 (Median) Sound (dBA) in 2000-2014 (below)

### Outcome

On the baseline and all subsequent biennial follow-up questionnaires, participants were asked to report all occurrences of clinician diagnosed CVD (coronary heart disease (CHD) or stroke) and participants (or next-of-kin for fatal cases) provided consent to review medical records pertaining to their diagnosis. Methods to confirm incident CVD in NHS have been published in detail elsewhere.^29–31^ In brief, incident CVD was determined as the first occurrence of either non-fatal myocardial infarction (MI) or fatal CHD (ICD-9 code 410) or non-fatal or fatal stroke (ICD-9 codes 430–437). Cases of non-fatal CVD were confirmed through medical record review or through interview or a letter confirming hospitalization for the non-fatal MI or stroke. Cases of fatal CHD or fatal stroke were confirmed through hospital record review, autopsy, report of the underlying cause on the death certificate, a history of CHD or stroke and CHD or stroke was the most plausible cause of death, or supporting information provided by a family member. Stroke sub-causes were classified according to the criteria of the National Survey of Stroke as due to ischemia (embolic or thrombotic; ICD-9 codes 430-432), hemorrhage (subarachnoid hemorrhage or intracerebral hemorrhage; ICD-9 codes 433-434 and 436), or other, unspecified/unknown cause.

### Exposure

National Park Service researchers created a geospatial noise model to predict outdoor sound levels for the contiguous US (**Figure 1**).^32, 33^ The model used acoustic data from 1.5 million hours of long-term measurements from 492 urban and rural monitoring sites located across the contiguous US during 2000-2014. The geospatial noise model regressed monitored sound-levels with spatial datasets of environmental factors such as topography, climate, hydrology, and anthropogenic activity. Using a random forest approach (a tree-based machine learning algorithm), the ensemble of spatial datasets that best predicted measured sound-levels were retained, and their relative contribution to sound-levels at monitoring sites was used to estimate sound levels in locations where monitoring stations were not present. The geospatial noise model was cross-validated using a leave-one-out cross-validation process. Cross-validated residuals relative to those from the null model indicate that root mean squared error ranged from 4.5-4.9 dB, median absolute deviation ranged from 2.3-2.4 dB, and the percent of variance explained ranged from 80-84% across the noise metrics that we considered. The resulting model enabled the mapping of sound levels at a 270 m x 270 m resolution across the contiguous US, over a 14-year period (long-term average). Noise estimates were appended to each residential address throughout follow-up (updated each time a participant moved address), based on the 270 m grid cell the address was contained within. We carried backwards and forwards the temporally invariant noise estimates from 2000-2014 to 1988-2018 addresses, making the assumption that relative noise levels did not change substantially over time.

Environmental sound levels vary diurnally and are summarized using a variety of statistics across multiple time scales and frequency ranges. Given the posited mechanism of nighttime noise to increase physiologic stress,^33^ coupled with the increased likelihood that participants were occupying their residence during nighttime hours, we used the anthropogenic nighttime (7pm-7am) A-weighted L50 sound pressure level metric as our primary noise exposure. The L50 is an exceedance metric that corresponds to the sound pressure level exceeded 50% of the time. A-weighting is an adjustment in decibels (dBA) that reflects how the human ear perceives sound across the frequency spectrum. While many noise measures are available from the National Park Service model, we focused on anthropogenic L50 nighttime (7pm-7am) and, for completeness, anthropogenic L50 daytime (7am-7pm) sound pressure levels in dBA.

### Statistical Analysis

Person-months of follow-up were accrued from the return date of the 1988 questionnaire until either the participant became a case, died, or the end of follow-up (May 31, 2018). We fit time-varying Cox proportional hazards regression models to calculate the hazard ratio (HR) and 95% confidence interval (95% CI) for developing CVD (CHD or stroke) associated with exposure to noise. We used restricted cubic splines to determine the linearity of exposure-response associations with a likelihood ratio test comparing the linear model to the model with linear and cubic spline terms. There were no deviations from linearity observed, therefore we present continuous exposure-response results, modeled per interquartile range (IQR). There were no violations of the proportional hazards assumption, which we tested by including interaction terms of noise exposure and age.

Models were stratified by age at follow-up and time period, and we examined the following covariates, determined *a priori* as potential confounders because they are CVD risk factors and/or may be correlated with noise (all variables are time-varying unless otherwise indicated): race (white/non-white, proxy for unmeasured consequences of racism, time invariant), smoking (current/past/never), pack-years smoked (continuous), family history of MI (yes/no), menopausal status (premenopausal/postmenopausal/dubious or missing), postmenopausal hormone use (premenopausal/never/current/former/missing), diet based on the Alternate Healthy Eating Index estimated from food frequency questionnaires^34^ (continuous), alcohol consumption (grams/day: 0/0.1-4.9/5.0-14.9/≥ 15/Missing), weight status (normal [body mass index (BMI) 18.5-24.9]/overweight [BMI 25-29.9]/obese [BMI > 30]), and night shift work before 1989 (never/1-14 years of shift work/15-29 years of shift work/≥30 years of shift work, time invariant). To account for individual socioeconomic status (SES), we included information on self-reported parental occupation for the participant’s mother (housewife/other, time invariant) and father (professional or manager/other, time invariant), whether the participant had a registered nursing degree (yes/no, time invariant), marital status (married/other), and husband’s highest educational attainment (< high school/high school graduate/>high school/missing or not married, time invariant). We accounted for area-level SES by using a neighborhood SES index that we developed specifically to differentiate neighborhood deprivation amongst NHS participants. We obtained the Census tract-level variables for the temporally closest Census from the Neighborhood Change Database (NCDB), which provides US Census data from 1970, 1980, 1990, 2000, and 2010 with normalized boundaries over time.^35^ To create the nSES score, we z-standardized and summed the following 9 variables: median household income, median home value, percent with a college degree, percent non-Hispanic White, percent non-Hispanic Black, percent of foreign-born residents, percent of families receiving interest or dividends, percent of occupied housing units, and percent unemployed.^36^ We classified Census tract-level population density as <1000 people per mile^2^ or ≥1000 people per mile^2^ and assigned Census region of the US (Northeast/Midwest/West/South). Fine particulate matter air pollution (PM2.5) estimates were linked to participants’ geocoded addresses; the air pollution exposure model is detailed elsewhere.^37^ Vegetation cover (greenness) in a 270 m buffer around each participant’s residential address was derived using Google Earth Engine from 30 m x 30 m Landsat Normalized Difference Vegetation Index (NDVI) data; the exposure assessment method is detailed elsewhere.^38^ Participant observations missing air pollution were excluded from analyses. The missing indicator method was used to account for missing categorical covariates.

To assess whether the association between noise exposure and CVD risk differed across susceptible subpopulations, we examined prespecified effect modification by population density (<1000 people per mile^2^/≥1000 people per mile^2^), region (Northeast/Midwest/West/South), air pollution quintile, greenness quintile, or nSES score quintile. To evaluate whether the association between continuous noise and CVD risk varied across levels of each potential modifier, we fit models that included a multiplicative interaction term between noise and the effect modifier. We reported stratum-specific HRs and 95% CIs and used likelihood ratio tests to determine statistical significance of departure from the null of no effect modification across levels of the modifier. In sensitivity analyses, we restricted analyses to 2000-2018 to assess the impact of carrying back the time-invariant noise estimates to 1988 (prior to noise sampling). We also assessed the impact of additionally adjusting for comorbidities (ever reported diabetes, elevated cholesterol, and/or high blood pressure), statin use, aspirin use, and physical activity (< 3 MET hours/week/3 to <9 MET hours/week/9 to <18 MET hours/week/≥ 18 MET hours/week) in fully adjusted models, as we believe these variables potentially lie along the causal pathway between noise and CVD.

We evaluated whether the relationship between noise exposure and CVD was mediated by sleep duration, which was self-reported by NHS participants on 1986, 2000, 2002, 2008, 2012, and 2014 questionnaires. We defined inadequate sleep duration as five hours or less per night, as in other NHS studies.^39^ We calculated the mediation proportion and its 95% CI by comparing the exposure effect estimate from the full model that includes the exposure, a potential intermediate variable, and any covariates with the exposure effect estimate obtained from a partial model that leaves out the intermediate variable. The mediation proportion is the proportion of increased CVD risk explained by higher exposure to noise that can be attributed to inadequate sleep. Confidence intervals for the mediation proportion were calculated using the data duplication method.^40^ Mediation analyses assumed that there was no unmeasured exposure–outcome confounding, no unmeasured mediator–outcome confounding, no unmeasured exposure–mediator confounding, and no mediator–outcome confounder affected by exposure. Although these assumptions are unverifiable, we included major confounders in our mediation analyses, and therefore, we believe our assumptions are reasonable. All analyses were conducted in SAS, version 9.4.

## Results

We observed 10,331 total CVD cases over 2,544,035 person-years of follow-up among the 114,116 eligible cohort members from 1988-2018. In cause-specific analyses, we observed 5,321 CHD cases and 5,010 stroke cases, of which, 1,929 were ischemic and 553 were hemorrhagic stroke, and 2,528 were unspecified. The mean age over follow-up was 67.6 years (**Table 1**). Participants were predominantly white and lived in the Northeastern US. Those living in areas with higher levels of nighttime noise lived in areas with higher population density, higher levels of air pollution, and were less likely to be white. Nighttime and daytime noise were correlated (0.73) and weakly correlated with other environmental variables (**Figure S1**).

**Table 1.** Age-Adjusted Nurses’ Health Study Participant Characteristics by Quintiles of Nighttime Anthropogenic L50 dBA from 1988-2018 (n = 114,116; averaged over 2,544,035 person-years)

| Characteristic | Total | Nighttime L<sub>50</sub> dBA Quintile 1 | Nighttime L<sub>50</sub> dBA Quintile 2 | Nighttime L<sub>50</sub> dBA Quintile 3 | Nighttime L<sub>50</sub> dBA Quintile 4 | Nighttime L<sub>50</sub> dBA Quintile 5 |
| --- | --- | --- | --- | --- | --- | --- |
|  | Mean (SD) or % | Mean (SD) or % | Mean (SD) or % | Mean (SD) or % | Mean (SD) or % | Mean (SD) or % |
| L<sub>50</sub> dBA anthropogenic nighttime | 43.0 (3.5) | 37.9 (3.5) | 41.9 (0.6) | 43.3 (0.4) | 44.7 (0.5) | 47.1 (1.3) |
| L<sub>50</sub> dBA anthropogenic daytime | 46.1 (4.2) | 40.8 (4.7) | 45.2 (2.4) | 47.1 (2.1) | 48.2 (2.1) | 49.3 (2.5) |
| Age (years)* | 67.6 (10.6) | 67.0 (10.7) | 67.3 (10.6) | 67.5 (10.6) | 67.9 (10.5) | 68.1 (10.5) |
| White (%) | 94 | 96 | 96 | 94 | 93 | 90 |
| Alternative Healthy Eating Index | 51.6 (10.2) | 51.6 (10.2) | 51.3 (10.1) | 51.6 (10.2) | 52.0 (10.2) | 51.8 (10.1) |
| Alcohol categories (%) |  |  |  |  |  |  |
| 0 grams/day | 29 | 30 | 29 | 28 | 27 | 29 |
| 0.1-4.9 grams/day | 19 | 19 | 19 | 19 | 19 | 18 |
| 5.0-14.9 grams/day | 12 | 12 | 12 | 13 | 12 | 11 |
| ≥ 15 grams/day | 8 | 8 | 8 | 8 | 8 | 7 |
| Missing | 32 | 30 | 31 | 32 | 33 | 35 |
| Body Mass Index categories (%) |  |  |  |  |  |  |
| Normal/Underweight | 41 | 41 | 41 | 42 | 42 | 39 |
| Overweight | 29 | 30 | 30 | 29 | 28 | 29 |
| Obese | 19 | 19 | 19 | 18 | 18 | 19 |
| Missing | 11 | 10 | 10 | 11 | 12 | 13 |
| Smoking Status (%) |  |  |  |  |  |  |
| Never smoker | 45 | 45 | 45 | 44 | 45 | 46 |
| Past smoker | 43 | 43 | 43 | 43 | 43 | 41 |
| Current smoker | 10 | 9 | 10 | 10 | 9 | 10 |
| Missing | 3 | 2 | 2 | 2 | 3 | 3 |
| Pack years of smoking | 11.7 (18.6) | 11.7 (18.5) | 11.8 (18.5) | 12.1 (18.9) | 11.6 (18.7) | 11.3 (18.6) |
| Have RN degree (%) | 74 | 76 | 75 | 74 | 73 | 72 |
| Married (%) | 63 | 66 | 65 | 64 | 62 | 60 |
| Husband's Highest Education (%) |  |  |  |  |  |  |
| < high school | 4 | 4 | 4 | 3 | 3 | 4 |
| high school grad | 26 | 29 | 28 | 25 | 23 | 25 |
| > high school education | 37 | 35 | 36 | 38 | 39 | 35 |
| Missing or not married | 34 | 31 | 32 | 33 | 35 | 37 |
| Housewife mother (%) | 64 | 63 | 64 | 64 | 64 | 64 |
| Professional or manager father, % | 26 | 25 | 25 | 26 | 27 | 25 |
| Family History of MI (%) | 26 | 26 | 27 | 26 | 26 | 25 |
| Menopausal status (%) |  |  |  |  |  |  |
| Premenopausal or missing | 44 | 46 | 44 | 44 | 43 | 43 |
| Postmenopausal | 56 | 54 | 56 | 56 | 57 | 57 |
| Postmenopausal hormone use (%) |  |  |  |  |  |  |
| Premenopausal | 8 | 8 | 8 | 8 | 8 | 8 |
| Never | 21 | 22 | 21 | 21 | 20 | 19 |
| Current | 22 | 21 | 21 | 22 | 23 | 23 |
| Former | 33 | 34 | 34 | 34 | 33 | 32 |
| Missing | 16 | 15 | 16 | 16 | 17 | 18 |
| Night Shift Work |  |  |  |  |  |  |
| Never (%) | 31 | 32 | 30 | 31 | 30 | 30 |
| 1-14 years shift work (%) | 39 | 39 | 40 | 39 | 39 | 37 |
| 15-29 years shift work (%) | 4 | 4 | 4 | 4 | 4 | 4 |
| 30+ years shift work (%) | 1 | 1 | 1 | 1 | 1 | 1 |
| Missing shift work (%) | 25 | 24 | 25 | 25 | 26 | 28 |
| Sleep <5 hours per night (%) | 6 | 6 | 6 | 6 | 6 | 7 |
| Cancer during follow-up (%) | 7 | 7 | 7 | 7 | 8 | 7 |
| Population density $\geq 1000$ per mile<sup>2</sup> (%) | 65 | 39 | 57 | 70 | 79 | 81 |
| Region (%) |  |  |  |  |  |  |
| Northeast | 45 | 58 | 55 | 46 | 37 | 31 |
| Midwest | 16 | 11 | 18 | 17 | 14 | 20 |
| South | 17 | 13 | 13 | 18 | 21 | 18 |
| West | 13 | 9 | 6 | 10 | 18 | 20 |
| 12-month average PM<sub>2.5</sub> ($\mu\text{g}/\text{m}^3$) | 12.6 (4.6) | 11.0 (4.1) | 12.3 (4.3) | 13.0 (4.5) | 13.2 (4.7) | 13.7 (5.0) |
| 12-month average PM<sub>2.5</sub> Quintile (%) |  |  |  |  |  |  |
| Quintile 1 | 20 | 35 | 19 | 15 | 16 | 15 |
| Quintile 2 | 20 | 28 | 25 | 18 | 15 | 14 |
| Quintile 3 | 20 | 18 | 23 | 22 | 20 | 18 |
| Quintile 4 | 20 | 12 | 19 | 23 | 25 | 23 |
| Quintile 5 | 20 | 7 | 15 | 22 | 25 | 31 |
| NDVI (in 270m distance buffer) | 0.36 (0.17) | 0.39 (0.18) | 0.38 (0.17) | 0.37 (0.16) | 0.35 (0.15) | 0.33 (0.15) |
| NDVI Quintile (%) |  |  |  |  |  |  |
| Quintile 1 | 18 | 16 | 15 | 16 | 19 | 26 |
| Quintile 2 | 19 | 16 | 17 | 19 | 20 | 21 |
| Quintile 3 | 19 | 17 | 19 | 21 | 20 | 19 |
| Quintile 4 | 19 | 19 | 21 | 21 | 19 | 16 |
| Quintile 5 | 19 | 27 | 22 | 19 | 16 | 13 |
| Neighborhood SES (time-varying) Quintile (%) |  |  |  |  |  |  |
| Quintile 1 | 20 | 31 | 24 | 17 | 14 | 16 |
| Quintile 2 | 21 | 20 | 23 | 20 | 18 | 22 |
| Quintile 3 | 23 | 21 | 22 | 23 | 22 | 24 |
| Quintile 4 | 13 | 11 | 12 | 13 | 15 | 15 |
| Quintile 5 | 23 | 17 | 19 | 26 | 31 | 22 |

\* Value is not age-adjusted

The associations between nighttime noise and CVD, CHD, and stroke (total or subtype-specific) risks are shown in **Table 2**. Each interquartile range increase in L50 nighttime noise (3.67 dBA) was associated with an HR of 1.04 (95% CI 1.02, 1.06) in fully adjusted models for CVD. Overall nighttime noise and CVD results were driven primarily by CHD (HR 1.05 [95% CI 1.02, 1.09]), as opposed to stroke (HR 1.02 [95% CI 0.99, 1.06]). Ischemic stroke and hemorrhagic stroke findings were not statistically significant, although there was a suggestive positive association of nighttime noise with hemorrhagic stroke (HR 1.08 [95% CI 0.98, 1.19]). Similar patterns were observed in analyses of L50 daytime noise.

**Table 2.** Hazard ratios (HR) and 95% confidence intervals (CIs) for anthropogenic L_50_ nighttime and anthropogenic L_50_ daytime sound pressure (dBA) and cardiovascular disease incidence in the Nurses’ Health Study from 1988 to 2018 (n = 114,116; person-years = 2,544,035). Total CVD (CHD or stroke), CHD, stroke, and stroke subtypes (ischemic and hemorrhagic stroke) are shown.

|  | L<sub>50</sub> nighttime noise |  | L<sub>50</sub> daytime noise |  |
| --- | --- | --- | --- | --- |
|  | Age and Calendar Year Adjusted HR (95% CI) | Fully Adjusted* HR (95% CI) | Age and Calendar Year Adjusted HR (95% CI) | Fully Adjusted* HR (95% CI) |
| **CVD (n = 10,331)** | 1.01 (0.99, 1.03) | 1.04 (1.02, 1.06) | 1.01 (0.99, 1.03) | 1.04 (1.02, 1.07) |
| **CHD (n = 5,321)** | 1.02 (1.00, 1.05) | 1.05 (1.02, 1.09) | 1.02 (0.99, 1.05) | 1.06 (1.02, 1.09) |
| **Stroke (n = 5,010)** | 1.00 (0.97, 1.02) | 1.02 (0.99, 1.06) | 1.00 (0.97, 1.03) | 1.03 (1.00, 1.06) |
| ***Ischemic stroke (n = 1,929)*** | 0.96 (0.92, 1.01) | 1.00 (0.96, 1.05) | 0.98 (0.93, 1.02) | 1.04 (0.99, 1.09) |
| ***Hemorrhagic stroke (n = 553)*** | 1.07 (0.98, 1.18) | 1.08 (0.98, 1.19) | 1.08 (0.99, 1.18) | 1.10 (0.99, 1.21) |

\* Hazard ratios are adjusted for age and calendar year, race, smoking status, pack-years smoked, family history of MI, menopausal status, postmenopausal hormone use, diet, alcohol consumption, BMI, night shift work, parental occupation, educational attainment (RN degree), marital status, husband's highest education, nSES score quintile, region, and air pollution (PM<sub>2.5</sub>). Note: IQR for anthropogenic L<sub>50</sub> nighttime sound: 3.67 dBA; IQR for anthropogenic L<sub>50</sub> daytime sound: 4.35 dBA.

Stratified analyses showed that associations of stroke with nighttime noise varied by population density (**Figure S2; Table S1**). We observed positive associations for stroke among those living in census tracts with <1000 people per mile^2^ and null association of nighttime noise with stroke among those living in census tracts with ≥1000 people per mile^2^ (p for interaction = 0.03). Nighttime and daytime noise associations with CVD outcomes did not significantly differ by all other prespecified effect modifiers (**Figure S3; Table S2**).

In sensitivity analyses, HR for nighttime and daytime noise with CVD were slightly smaller in prespecified analyses restricted to 2000-2018 follow-up (**Table S3; Table S4**). We found no appreciable difference in associations when time-varying statin use or aspirin use was added to fully adjusted models (**Table S5; Table S6**). The impact of adjustment for physical activity in fully adjusted models, which may be considered a mediator of the association between noise exposure and cardiometabolic diseases,^41^ resulted in no change in associations. Finally, additionally adjusting models for population density resulted in slightly stronger associations of nighttime (HR 1.06 [95% CI 1.03, 1.09]) and daytime (HR 1.07 [95% CI 1.03, 1.10]) noise with CHD, however, we chose not to adjust for population density in our main fully-adjusted model as it was a predictor used in NPS modelling of noise exposure.

Mediation analyses showed no evidence that the association of nighttime noise and CVD risk was mediated by inadequate sleep duration, defined as self-reported five hours or less of sleep per night (**Table S7**).

## Discussion

In this nationwide prospective analysis of female nurses, we observed small positive associations of nighttime and daytime noise exposure with total CVD (nighttime HR 1.04 [95% CI 1.02, 1.06]; daytime HR 1.04 [95% CI 1.02, 1.07]) that were robust to adjustment for important CVD risk factors. CVD associations were slightly stronger for CHD than CVD (CHD and stroke, combined). Associations of noise with CVD did not substantially differ by prespecified effect modifiers. We did not observe evidence that the relationship between noise and CVD risk was mediated by self-inadequate sleep duration.

Our findings were generally consistent with previous analyses of environmental noise exposure and CVD. In 2018, the WHO stated that high-quality and moderate-quality evidence was available to conclude that road traffic noise increased the risk of CHD and stroke, respectively.^24^ Since the 2018 WHO publication, evidence on noise and stroke has shown mixed findings and a 2021 publication called for further assessment of noise and stroke subtypes, specifically ischemic stroke, using richly-contextualized, individual-level longitudinal studies.^4^ We did not find evidence of an association of noise and ischemic stroke in the NHS. In agreement with other cohort studies, our study found increased risk of CHD (non-fatal MI or fatal CHD) associated with noise, but small or insignificant associations for stroke or stroke-subtypes.^22, 23, 42^ Similarly, a UK Biobank cohort study found suggestive positive associations of traffic noise with acute coronary events (including MI), after adjustment for confounders (HR 1.03 [95% CI 0.99, 1.08]), but found no evidence for stroke or stroke subtypes.^22^

An analysis of 2009 Medicare data on adults older than 65 living around airports in the US linked to Federal Aviation Administration aircraft noise contour data found that a 10 dBA higher aircraft noise exposure at the zip code level was associated with higher cardiovascular hospital admission rate (3.5% [95% CI 0.2%, 7.0%]), after controlling for age, sex, race, zip code level socioeconomic status and demographics, zip code level air pollution (PM2.5 and ozone), and roadway density.^43^ Additionally, a 2022 US study examined the relationship of airport noise at airport-adjacent residential address and hypertension, an important risk factor for CHD and stroke, and found suggestive evidence of a positive association.^44^ Our study is the first to provide longitudinal, individual-level evidence of associations of anthropogenic noise (from multiple sources including traffic, aircraft, industry, etc.) with total CVD (CHD and stroke), CHD, and stroke.

There is no reason to believe that the biological pathways mediating effects of noise on human health differ in US citizens compared to those from other countries, however, US-based evidence is crucial. Although associations of CVD with anthropogenic noise in our study were small, a 4% increased risk per interquartile range increase could be an important long term risk factor for CVD because such a large percentage of the population is exposed. In addition, this 4% increase is likely an underestimate due to potential non-differential exposure measurement error. More refined noise exposure models with higher spatial resolution in urban areas (i.e., with greater exposure contrast among addresses in loudest areas) may result in stronger associations with CVD outcomes. Indeed, a Swiss cohort study showed that reducing exposure measurement error by modelling traffic noise using high spatiotemporal resolution inputs compared to lower resolution inputs resulted in stronger associations with MI.^45^ US noise models could be improved by using sound propagation models that incorporate reflection, diffraction, and absorption by sound barriers (e.g. buildings).^15, 18, 46^ Development of a US source-specific (e.g., traffic) noise model with high spatial resolution inputs will be important in future analyses for accurately estimating health effects of noise and providing policy-relevant evidence.

This study has limitations. Firstly, the nationwide noise model approximated outdoor noise exposure at a 270 m x 270 m resolution, which may not accurately capture noise variability over fine spatial scales, particularly in urban environments that have high variability in emissions, reflection, diffraction, and absorption.^45^ While our predictions of outdoor noise contained measurement error, the noise model performed well in cross-validation (R^2^≥0.8).^32^ In addition, while information on changes in residential address were incorporated over follow-up, the noise model surface did not vary over time. Due to using long-term average noise predictions (2000-2014), we back-extrapolated the time-invariant exposure to biennially updated addresses between 1988-1999, which could result in exposure misclassification; however, in sensitivity analyses we found similar associations with slightly larger confidence intervals when using 2000 as a baseline, potentially arising from substantial participant exclusions due to prior CVD before 2000. Another limitation is that we could not measure factors that might alter individual noise exposure, such as housing quality or noise in the indoor environment. Also, our noise model did not differentiate between specific sources of sound, including road traffic, airports, and industrial land uses. For these reasons, there was likely unsystematic, nondifferential error in measurement of true exposure to noise which may bias our observed associations towards the null.

With any study of neighborhood factors and health, there is a possibility that participants may self-select into certain neighborhoods they deem healthier than others. Therefore, healthier individuals may have chosen to move to neighborhoods with lower levels of noise, which may explain our findings. However, adjustment for multiple CVD risk factors along with our prospective analysis of incident CVD would decrease the potential for this type of bias and a study has demonstrated residential self-selection is not a major concern in NHS.^47^ We did not find evidence of effect mediation by short sleep duration, however, humans react to environmental sounds while asleep^3, 48^ and, irrespective of sleep impairment and cognitive perception of noise,^49^ nighttime noise has been shown in experimental studies to induce both endothelial dysfunction and prothrombotic inflammatory changes to the plasma proteome in healthy individuals.^50, 51^ Therefore self-reported short sleep duration may not adequately capture the breadth of potential effects of nighttime noise on CVD risk. Because all participants were female nurses at enrollment, the generalizability of our findings to males and lower SES, non-working groups is also potentially limited. Finally, although we adjusted for numerous risk factors for CVD, we cannot rule out the possibility that other factors that are correlated with noise might explain the observed association between noise exposure and CVD risk.

This analysis has several strengths. Firstly, this nationwide, 30-year prospective analysis included residence-level metrics of exposure to nighttime and daytime noise with medical record confirmed or participant/medical professional corroborated CVD endpoints. Secondly, our analyses incorporated time-varying individual-level information on important CVD risk factors, including smoking, family history of MI, diet, menopausal status, postmenopausal hormone use, BMI, night shift work, as well as area-level data on neighborhood SES, region, and residential greenness, and air pollution. Because of the breadth of information available in the NHS cohort, we were able to conduct stratified analyses, as well as to examine potential mediation of the noise and CVD relationship by short sleep duration. Finally, this nationwide study covered a broad geographic region with a considerable range of noise levels and included participants residing in high and low population density areas of the US, which increases the generalizability of findings. A strength of the noise model used in this study was the extensive noise monitoring in rural areas, near national parks,^52^ which allowed for well-validated prediction of noise in rural locations and, in turn, a nationwide analysis that included predictions of noise at low levels. This contrasts the noise models used in European epidemiological analyses, which assign a standard minimum noise level (e.g. 20dBA) due to limited monitoring and/or traffic data availability in rural areas.^15, 46^

This prospective study, conducted over 30 years of follow-up with objective measures of anthropogenic nighttime and daytime noise across the entire US, provides evidence that nighttime noise was modestly associated with CVD incidence and nighttime and daytime noise was associated with incident CHD, after accounting for individual and area-level risk factors for CVD.

## Data Availability

Access to Channing Division of Network Medicine Cohorts, which include the Nurses Health Study, requires the proposal and approval of the specific research project and is not sharable.

## Supplementary material

This supplement provides results from: correlation (Figure S1); stratified analyses that correspond to Figure S2 and S3 (Tables S1 and S2); time-restricted Table 1 and Cox PH analyses with baseline in 2000 (Tables S3 and S4, respectively); and additional Table 1 variables for sensitivity analyses and Cox PH sensitivity analyses (Tables S5 and S6); mediation analyses (Table S7).

**Figure S1.**
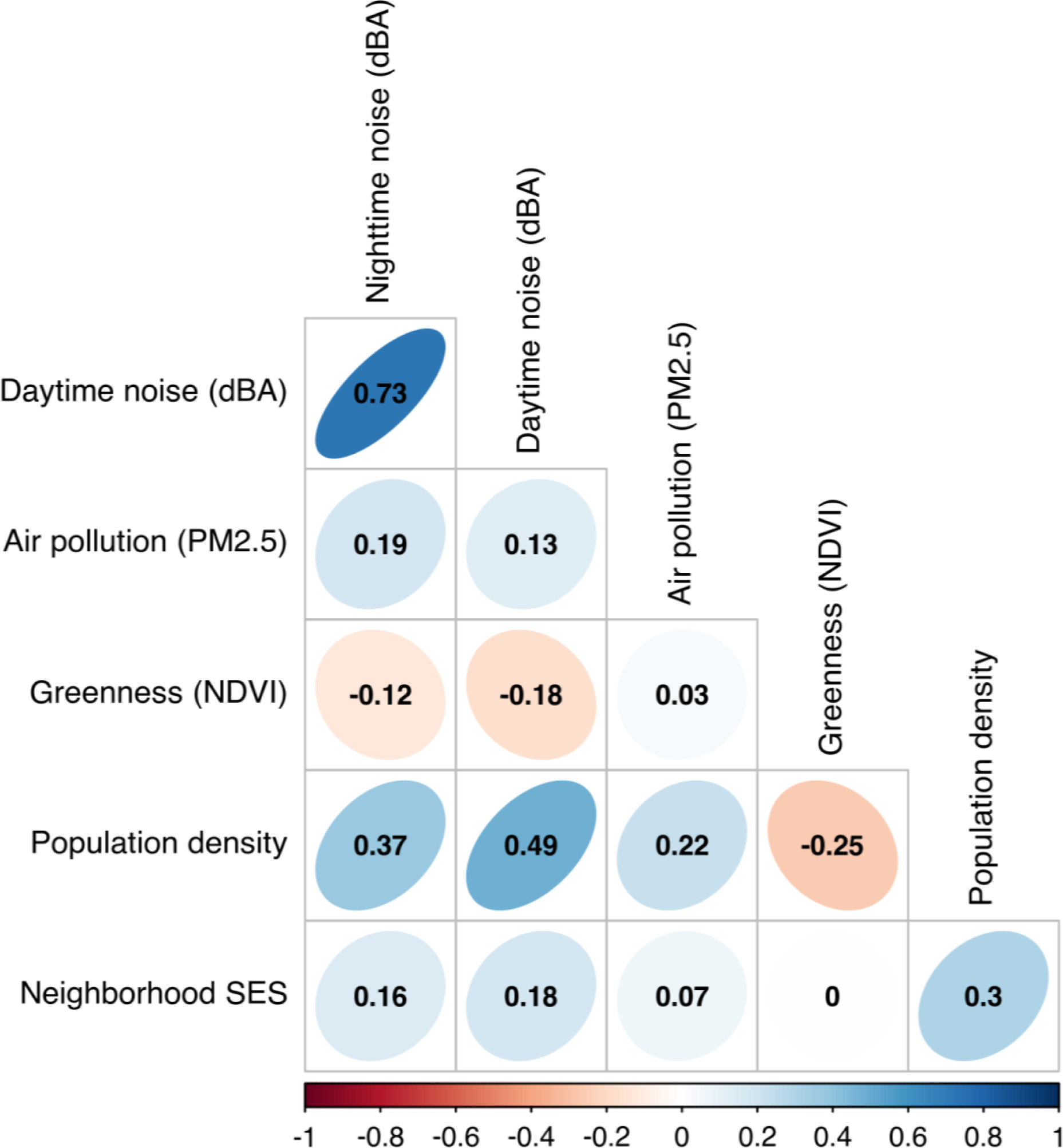
Spearman’s rank correlation plot of nighttime anthropogenic L_50_ (median) sound (dBA), daytime anthropogenic L_50_ (median) sound (dBA), air pollution (PM_2.5_), greenness (NDVI), population density (Census population per square mile), and neighborhood socioeconomic status (z-score)

**Figure 2.**
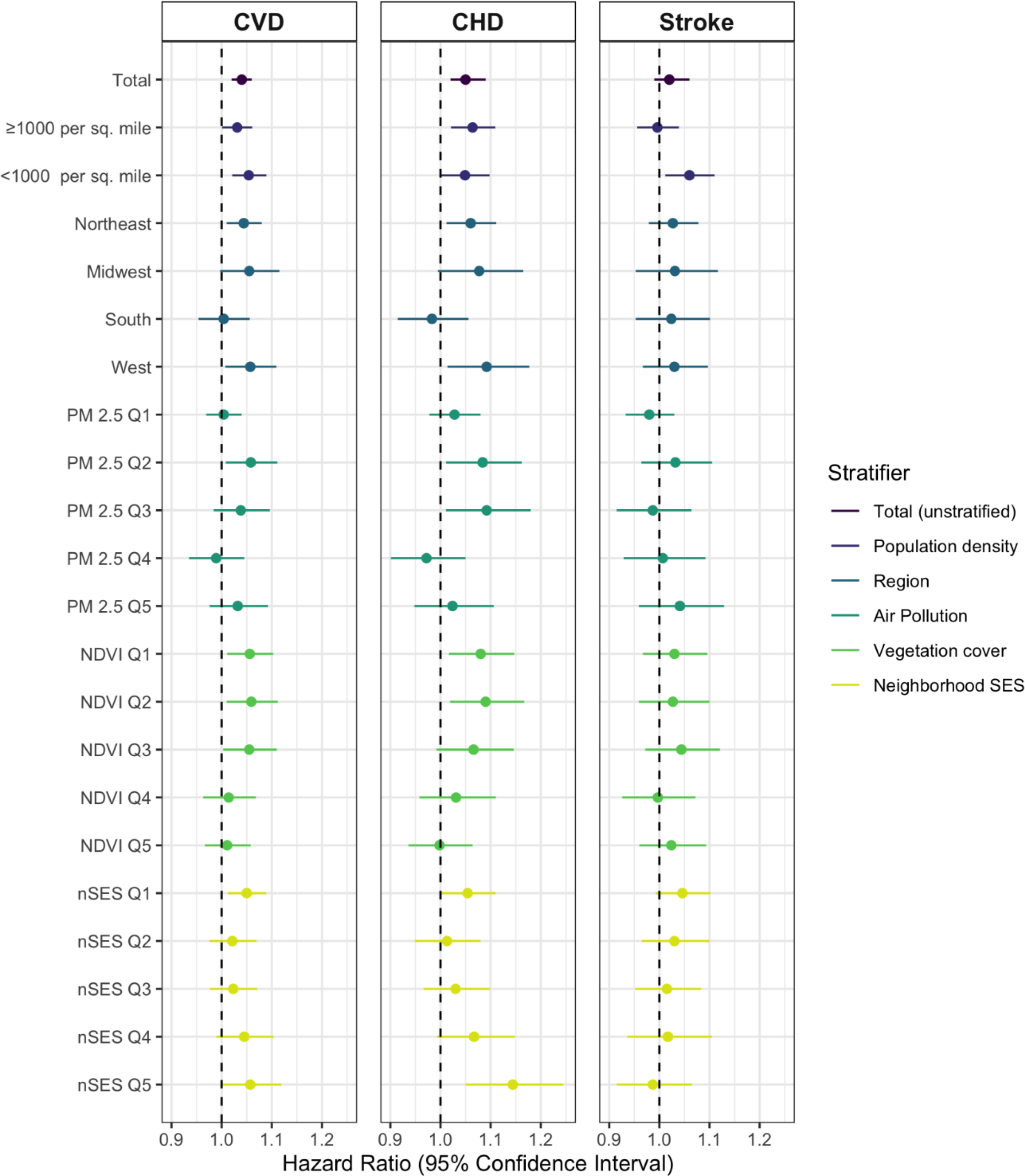
Stratified Analyses of Anthropogenic Nighttime L_50_ (Median) Sound (dBA) for CVD, CHD, and Stroke incidence in the Nurses’ Health Study (n = 114,116; 1988-2018) ^a^ Hazard ratios are adjusted for age and calendar year, race, smoking status, pack-years smoked, family history of MI, menopausal status, postmenopausal hormone use, diet, alcohol consumption, BMI, night shift work, parental occupation, educational attainment (RN degree), marital status, husband’s highest education, neighborhood SES score, region, and air pollution (PM2.5). Stratifying variables are not adjusted in respective analyses. Note: IQR for anthropogenic L50 nighttime sound: 3.67 dBA.

**Figure 3.**
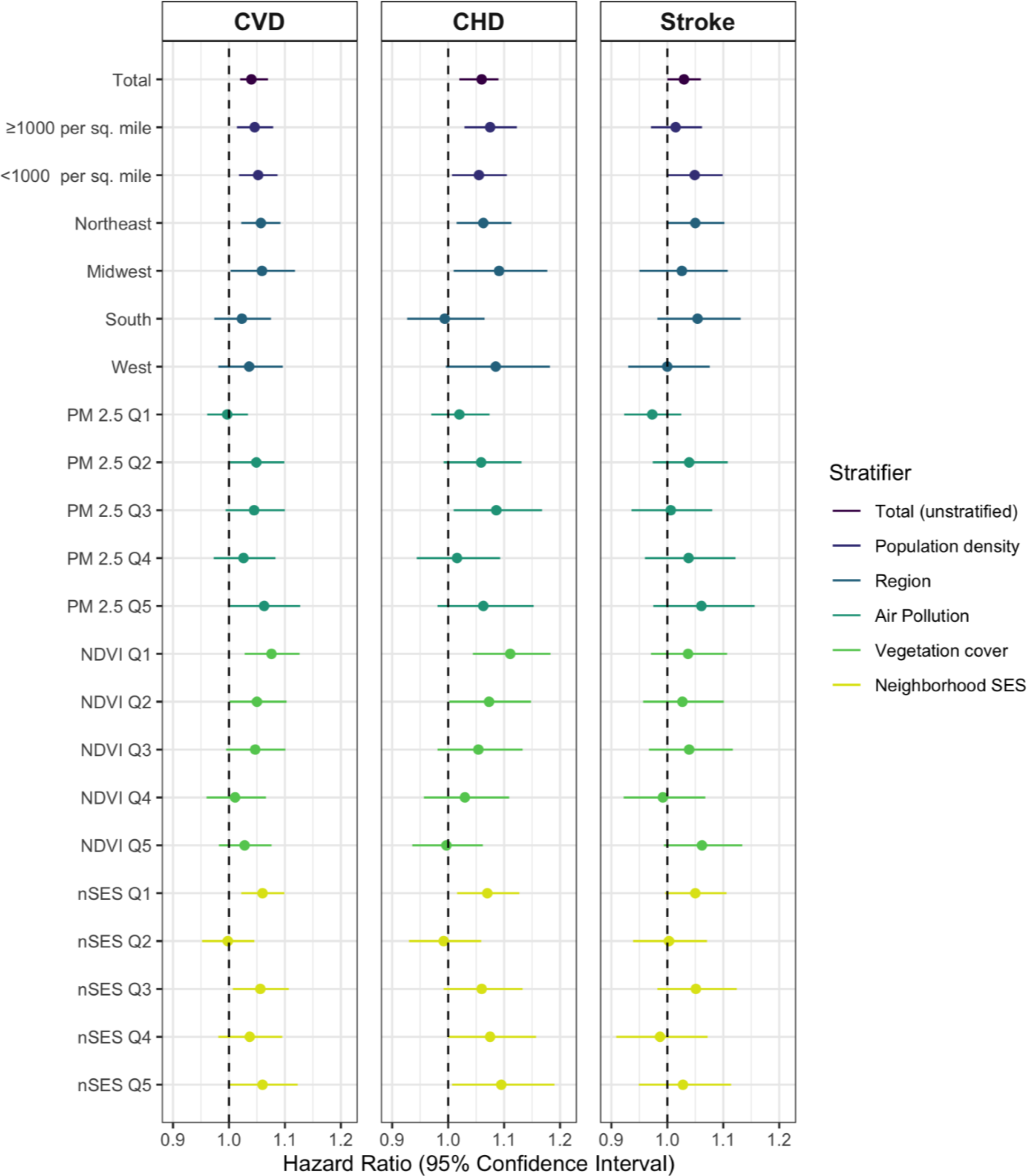
Stratified Analyses of Anthropogenic Daytime L_50_ (Median) Sound (dBA) for CVD, CHD, and Stroke incidence in the Nurses’ Health Study (n = 114,116; 1988-2018) ^a^ Hazard ratios are adjusted for age and calendar year, race, smoking status, pack-years smoked, family history of MI, menopausal status, postmenopausal hormone use, diet, alcohol consumption, BMI, night shift work, parental occupation, educational attainment (RN degree), marital status, husband’s highest education, neighborhood SES score, region, and air pollution (PM2.5). Stratifying variables are not adjusted in respective analyses. Note: IQR for anthropogenic L50 daytime sound: 4.35 dBA.

**Figure S2. Stratified Analyses of Anthropogenic Nighttime L_50_ (Median) Sound (dBA) for CVD, CHD, and Stroke incidence in the Nurses’ Health Study (n = 114,116; 1988-2018).**

**Figure S3. Stratified Analyses of Anthropogenic Daytime L50 (Median) Sound (dBA) for CVD, CHD, and Stroke incidence in the Nurses’ Health Study (n = 114,116; 1988-2018).**

**Table S1.** Nurses’ Health Study Person-time, CVD, CHD, stroke cases and fully adjusted HR and 95% confidence intervals for nighttime anthropogenic L_50_ (median) sound (dBA) from 1988-2018 (n = 114,116) by stratifying variables (population density, region, air pollution, greenness, and neighborhood socio-economic status)

|  | Person-time | CVD cases | CHD cases | Stroke cases | CVD Fully Adjusted<sup>a</sup> HR (95% CI) | CHD Fully Adjusted<sup>a</sup> HR (95% CI) | Stroke Fully Adjusted<sup>a</sup> HR (95% CI) |
| --- | --- | --- | --- | --- | --- | --- | --- |
| Total (unstratified) | 2,544,035 | 10,331 | 5,321 | 5,010 | 1.04 (1.02, 1.06) | 1.05 (1.02, 1.09) | 1.02 (0.99, 1.06) |
| Population density |  |  |  |  |  |  |  |
| ≥1000 people per mile<sup>2</sup> | 1,652,688 | 6,828 | 3,548 | 3,280 | 1.03 (1.00, 1.06) | 1.06 (1.02, 1.11) | 1.00 (0.96, 1.04) |
| <1000 people per mile<sup>2</sup> | 891,348 | 3,503 | 1,773 | 1,730 | 1.05 (1.02, 1.09) | 1.05 (1.00, 1.10) | 1.06 (1.01, 1.11) |
| *p-for-interaction* |  |  |  |  | 0.17 | 0.54 | 0.03 |
| Region |  |  |  |  |  |  |  |
| Northeast | 1,157,696 | 4,457 | 2,320 | 2,137 | 1.04 (1.01, 1.08) | 1.06 (1.01, 1.11) | 1.03 (0.98, 1.08) |
| Midwest | 405,844 | 1,582 | 815 | 767 | 1.05 (1.00, 1.12) | 1.08 (0.99, 1.17) | 1.03 (0.95, 1.12) |
| South | 421,149 | 1,692 | 831 | 861 | 1.00 (0.95, 1.06) | 0.98 (0.92, 1.06) | 1.02 (0.95, 1.10) |
| West | 319,628 | 1,345 | 607 | 738 | 1.06 (1.01, 1.11) | 1.09 (1.01, 1.18) | 1.03 (0.97, 1.10) |
| *p-for-interaction* |  |  |  |  | 0.25 | 0.16 | 0.48 |
| Air pollution PM2.5 |  |  |  |  |  |  |  |
| Quintile 1 | 508,998 | 2,066 | 1,082 | 984 | 1.00 (0.97, 1.04) | 1.03 (0.98, 1.08) | 0.98 (0.93, 1.03) |
| Quintile 2 | 510,635 | 2,027 | 1,009 | 1,018 | 1.06 (1.01, 1.11) | 1.08 (1.01, 1.16) | 1.03 (0.96, 1.10) |
| Quintile 3 | 510,407 | 2,021 | 1,021 | 1,000 | 1.04 (0.98, 1.10) | 1.09 (1.01, 1.18) | 0.99 (0.92, 1.06) |
| Quintile 4 | 509,846 | 2,063 | 1,079 | 984 | 0.99 (0.94, 1.05) | 0.97 (0.90, 1.05) | 1.01 (0.93, 1.09) |
| Quintile 5 | 504,151 | 2,154 | 1,130 | 1,024 | 1.03 (0.98, 1.09) | 1.02 (0.95, 1.11) | 1.04 (0.96, 1.13) |
| *p-for-interaction* |  |  |  |  | 0.22 | 0.16 | 0.48 |
| Greenness NDVI |  |  |  |  |  |  |  |
| Quintile 1 | 470,012 | 2,069 | 1,112 | 957 | 1.06 (1.01, 1.10) | 1.08 (1.02, 1.15) | 1.03 (0.97, 1.10) |
| Quintile 2 | 477,625 | 2,075 | 1,096 | 979 | 1.06 (1.01, 1.11) | 1.09 (1.02, 1.17) | 1.03 (0.96, 1.10) |
| Quintile 3 | 482,536 | 2,024 | 1,017 | 1,007 | 1.06 (1.00, 1.11) | 1.07 (0.99, 1.15) | 1.04 (0.97, 1.12) |
| Quintile 4 | 487,865 | 1,825 | 930 | 895 | 1.01 (0.96, 1.07) | 1.03 (0.96, 1.11) | 1.00 (0.93, 1.07) |
| Quintile 5 | 491,496 | 1,852 | 933 | 919 | 1.01 (0.97, 1.06) | 1.00 (0.94, 1.06) | 1.02 (0.96, 1.09) |
| *p-for-interaction* |  |  |  |  | 0.30 | 0.28 | 0.75 |
| Time-varying nSES score |  |  |  |  |  |  |  |
| Quintile 1 | 520,565 | 2,472 | 1,254 | 1,218 | 1.05 (1.01, 1.09) | 1.05 (1.00, 1.11) | 1.05 (0.99, 1.10) |
| Quintile 2 | 527,630 | 2,203 | 1,118 | 1,085 | 1.02 (0.98, 1.07) | 1.01 (0.95, 1.08) | 1.03 (0.97, 1.10) |
| Quintile 3 | 572,052 | 2,134 | 1,103 | 1,031 | 1.02 (0.98, 1.07) | 1.03 (0.97, 1.10) | 1.02 (0.95, 1.08) |
| Quintile 4 | 334,704 | 1,516 | 880 | 636 | 1.05 (0.99, 1.10) | 1.07 (0.99, 1.15) | 1.02 (0.94, 1.10) |
| Quintile 5 | 589,084 | 2,006 | 966 | 1,040 | 1.06 (1.00, 1.12) | 1.14 (1.05, 1.25) | 0.99 (0.92, 1.07) |
| *p-for-interaction* |  |  |  |  | 0.65 | 0.21 | 0.61 |

<sup>a</sup> Hazard ratios are adjusted for age and calendar year, race, smoking status, pack-years smoked, family history of MI, menopausal status, postmenopausal hormone use, diet, alcohol consumption, BMI, night shift work, parental occupation, educational attainment (RN degree), marital status, husband's highest education, neighborhood SES score, region, and air pollution (PM<sub>2.5</sub>). Stratifying variables are not adjusted in respective analyses. Note: IQR for anthropogenic L<sub>50</sub> nighttime sound: 3.67 dBA.

**Table S2.** Nurses’ Health Study Person-time, CVD, CHD, stroke cases and fully adjusted HR and 95% confidence intervals for daytime anthropogenic L_50_ (median) sound (dBA) from 1988-2018 (n = 114,116) by stratifying variables (population density, region, air pollution, greenness, and neighborhood socio-economic status)

|  | Person-time | CVD cases | CHD cases | Stroke cases | CVD Fully Adjusted<sup>a</sup> HR (95% CI) | CHD Fully Adjusted<sup>a</sup> HR (95% CI) | Stroke Fully Adjusted<sup>a</sup> HR (95% CI) |
| --- | --- | --- | --- | --- | --- | --- | --- |
| Total (unstratified) | 2,544,035 | 10,331 | 5,321 | 5,010 | 1.04 (1.02, 1.07) | 1.06 (1.02, 1.09) | 1.03 (1.00, 1.06) |
| Population density |  |  |  |  |  |  |  |
| ≥1000 people per mile<sup>2</sup> | 1,652,688 | 6,828 | 3,548 | 3,280 | 1.05 (1.01, 1.08) | 1.07 (1.03, 1.12) | 1.02 (0.97, 1.06) |
| <1000 people per mile<sup>2</sup> | 891,348 | 3,503 | 1,773 | 1,730 | 1.05 (1.02, 1.09) | 1.05 (1.01, 1.10) | 1.05 (1.00, 1.10) |
| *p-for-interaction* |  |  |  |  | 0.35 | 0.45 | 0.15 |
| Region |  |  |  |  |  |  |  |
| Northeast | 1,157,696 | 4,457 | 2,320 | 2,137 | 1.06 (1.02, 1.09) | 1.06 (1.01, 1.11) | 1.05 (1.00, 1.10) |
| Midwest | 405,844 | 1,582 | 815 | 767 | 1.06 (1.00, 1.12) | 1.09 (1.01, 1.18) | 1.03 (0.95, 1.11) |
| South | 421,149 | 1,692 | 831 | 861 | 1.02 (0.97, 1.08) | 0.99 (0.93, 1.07) | 1.05 (0.98, 1.13) |
| West | 319,628 | 1,345 | 607 | 738 | 1.04 (0.98, 1.10) | 1.09 (1.00, 1.18) | 1.00 (0.93, 1.08) |
| *p-for-interaction* |  |  |  |  | 0.39 | 0.22 | 0.27 |
| Air pollution PM2.5 |  |  |  |  |  |  |  |
| Quintile 1 | 508,998 | 2,066 | 1,082 | 984 | 1.00 (0.96, 1.03) | 1.02 (0.97, 1.07) | 0.97 (0.92, 1.03) |
| Quintile 2 | 510,635 | 2,027 | 1,009 | 1,018 | 1.05 (1.00, 1.10) | 1.06 (0.99, 1.13) | 1.04 (0.97, 1.11) |
| Quintile 3 | 510,407 | 2,021 | 1,021 | 1,000 | 1.05 (0.99, 1.10) | 1.09 (1.01, 1.17) | 1.01 (0.94, 1.08) |
| Quintile 4 | 509,846 | 2,063 | 1,079 | 984 | 1.03 (0.97, 1.08) | 1.02 (0.94, 1.09) | 1.04 (0.96, 1.12) |
| Quintile 5 | 504,151 | 2,154 | 1,130 | 1,024 | 1.06 (1.00, 1.13) | 1.06 (0.98, 1.15) | 1.06 (0.97, 1.16) |
| *p-for-interaction* |  |  |  |  | 0.19 | 0.56 | 0.24 |
| Greenness NDVI |  |  |  |  |  |  |  |
| Quintile 1 | 470,012 | 2,069 | 1,112 | 957 | 1.08 (1.03, 1.13) | 1.11 (1.04, 1.18) | 1.04 (0.97, 1.11) |
| Quintile 2 | 477,625 | 2,075 | 1,096 | 979 | 1.05 (1.00, 1.10) | 1.07 (1.00, 1.15) | 1.03 (0.96, 1.10) |
| Quintile 3 | 482,536 | 2,024 | 1,017 | 1,007 | 1.05 (0.99, 1.10) | 1.05 (0.98, 1.13) | 1.04 (0.97, 1.12) |
| Quintile 4 | 487,865 | 1,825 | 930 | 895 | 1.01 (0.96, 1.07) | 1.03 (0.96, 1.11) | 0.99 (0.92, 1.07) |
| Quintile 5 | 491,496 | 1,852 | 933 | 919 | 1.03 (0.98, 1.08) | 1.00 (0.94, 1.06) | 1.06 (0.99, 1.13) |
| *p-for-interaction* |  |  |  |  | 0.30 | 0.15 | 0.55 |
| Time-varying nSES score |  |  |  |  |  |  |  |
| Quintile 1 | 520,565 | 2,472 | 1,254 | 1,218 | 1.06 (1.02, 1.10) | 1.07 (1.02, 1.13) | 1.05 (1.00, 1.11) |
| Quintile 2 | 527,630 | 2,203 | 1,118 | 1,085 | 1.00 (0.95, 1.04) | 0.99 (0.93, 1.06) | 1.00 (0.94, 1.07) |
| Quintile 3 | 572,052 | 2,134 | 1,103 | 1,031 | 1.06 (1.01, 1.11) | 1.06 (0.99, 1.13) | 1.05 (0.98, 1.12) |
| Quintile 4 | 334,704 | 1,516 | 880 | 636 | 1.04 (0.98, 1.09) | 1.08 (1.00, 1.16) | 0.99 (0.91, 1.07) |
| Quintile 5 | 589,084 | 2,006 | 966 | 1,040 | 1.06 (1.00, 1.12) | 1.09 (1.01, 1.19) | 1.03 (0.95, 1.11) |
| *p-for-interaction* |  |  |  |  | 0.22 | 0.30 | 0.47 |

\*486 CVD cases (233 CHD cases, 253 stroke cases) were missing NDVI; 1,255 CVD cases (748 CHD cases, 507 stroke cases) were missing region.
<sup>a</sup> Hazard ratios are adjusted for age and calendar year, race, smoking status, pack-years smoked, family history of MI, menopausal status, postmenopausal hormone use, diet, alcohol consumption, BMI, night shift work, parental occupation, educational attainment (RN degree), marital status, husband's highest education, neighborhood SES score, region, and air pollution (PM<sub>2.5</sub>). Stratifying variables are not adjusted in respective analyses. Note: IQR for anthropogenic L<sub>50</sub> daytime sound: 4.35 dBA.

**Table S3.** Age-Adjusted Nurses’ Health Study Participant Characteristics by Quintiles of Nighttime Anthropogenic L_50_ dBA from 2000-2018 (n = 95,225)

| Characteristic | Total | Nighttime L<sub>50</sub><br>dBA<br>Quintile 1 | Nighttime L<sub>50</sub><br>dBA<br>Quintile 2 | Nighttime L<sub>50</sub><br>dBA<br>Quintile 3 | Nighttime L<sub>50</sub><br>dBA<br>Quintile 4 | Nighttime L<sub>50</sub><br>dBA<br>Quintile 5 |
| --- | --- | --- | --- | --- | --- | --- |
|  | Mean (SD)<br>or % | Mean (SD)<br>or % | Mean (SD)<br>or % | Mean (SD)<br>or % | Mean (SD)<br>or % | Mean (SD)<br>or % |
| L<sub>50</sub> dBA anthropogenic nighttime | 43.1 (3.43) | 37.2 (3.65) | 41.7 (0.54) | 43.2 (0.39) | 44.6 (0.46) | 47.0 (1.28) |
| L<sub>50</sub> dBA anthropogenic daytime | 46.2 (4.08) | 40.2 (4.86) | 44.8 (2.46) | 46.8 (2.17) | 48.0 (2.09) | 49.1 (2.52) |
| Age (years)<sup>a</sup> | 73.4 (8.12) | 72.2 (7.96) | 72.6 (8.1) | 72.8 (8.09) | 73.2 (8.18) | 73.4 (8.21) |
| White (%) | 94 | 96 | 96 | 95 | 92 | 90 |
| Alternative Healthy Eating Index | 53.6 (10.4) | 53.4 (10.3) | 53.0 (10.1) | 53.3 (10.4) | 53.6 (10.3) | 53.4 (10.1) |
| Alcohol categories (%) |  |  |  |  |  |  |
| 0 grams/day | 29 | 29 | 29 | 28 | 27 | 28 |
| 0.1-4.9 grams/day | 17 | 16 | 17 | 16 | 16 | 15 |
| 5.0-14.9 grams/day | 12 | 12 | 12 | 12 | 12 | 10 |
| ≥ 15 grams/day | 9 | 10 | 9 | 9 | 8 | 8 |
| Missing | 33 | 33 | 35 | 35 | 37 | 39 |
| Body Mass Index categories (%) |  |  |  |  |  |  |
| Normal/Underweight | 40 | 40 | 39 | 40 | 40 | 37 |
| Overweight | 29 | 30 | 30 | 29 | 28 | 28 |
| Obese | 19 | 20 | 20 | 19 | 18 | 19 |
| Missing | 11 | 11 | 12 | 12 | 14 | 16 |
| Smoking Status (%) |  |  |  |  |  |  |
| Never smoker | 46 | 46 | 45 | 44 | 45 | 47 |
| Past smoker | 46 | 45 | 45 | 46 | 45 | 43 |
| Current smoker | 6 | 6 | 6 | 7 | 6 | 6 |
| Missing | 3 | 3 | 3 | 3 | 4 | 4 |
| Pack years of smoking | 11.4 (18.7) | 11.2 (18.5) | 11.4 (18.4) | 11.8 (19.0) | 11.2 (18.8) | 10.9 (18.6) |
| Have RN degree (%) | 74 | 75 | 73 | 73 | 71 | 69 |
| Married (%) | 56 | 61 | 57 | 56 | 53 | 49 |
| Husband's Highest Education (%) |  |  |  |  |  |  |
| < high school | 3 | 4 | 4 | 3 | 3 | 3 |
| high school grad | 26 | 28 | 27 | 25 | 22 | 23 |
| > high school education | 38 | 36 | 36 | 38 | 38 | 35 |
| Missing or not married | 33 | 32 | 34 | 34 | 37 | 39 |
| Housewife mother (%) | 64 | 63 | 64 | 63 | 64 | 64 |
| Professional or manager father, % | 26 | 26 | 25 | 27 | 27 | 26 |
| Family History of MI (%) | 25 | 24 | 25 | 25 | 25 | 24 |
| Menopausal status (%) |  |  |  |  |  |  |
| Premenopausal or missing | 64 | 62 | 61 | 61 | 61 | 60 |
| Postmenopausal | 36 | 38 | 39 | 39 | 39 | 40 |
| Postmenopausal hormone use (%) |  |  |  |  |  |  |
| Premenopausal | 1 | 1 | 1 | 1 | 1 | 1 |
| Never | 18 | 19 | 18 | 18 | 17 | 16 |
| Current | 15 | 16 | 15 | 16 | 16 | 16 |
| Former | 47 | 46 | 46 | 45 | 44 | 43 |
| Missing | 20 | 18 | 20 | 20 | 22 | 24 |
| Night Shift Work |  |  |  |  |  |  |
| Never (%) | 31 | 32 | 30 | 31 | 29 | 29 |
| 1-14 years shift work (%) | 39 | 39 | 39 | 38 | 38 | 36 |
| 15-29 years shift work (%) | 4 | 4 | 4 | 4 | 3 | 4 |
| 30+ years shift work (%) | 1 | 1 | 1 | 1 | 1 | 1 |
| Missing shift work (%) | 25 | 25 | 26 | 27 | 28 | 30 |
| Sleep <5 hours per night (%) | 7 | 7 | 7 | 7 | 7 | 7 |
| Cancer during follow-up (%) | 9 | 9 | 9 | 9 | 9 | 9 |
| Population density $\geq 1000$ per mile<sup>2</sup> (%) | 64 | 29 | 55 | 74 | 81 | 85 |
| 12-month average PM<sub>2.5</sub> ($\mu\text{g}/\text{m}^3$) | 11.32 (3.8) | 9.26 (3.19) | 11.0 (3.49) | 11.7 (3.66) | 12.1 (3.77) | 12.7 (3.96) |
| 12-month average PM<sub>2.5</sub> Quintile (%) |  |  |  |  |  |  |
| Quintile 1 | 20 | 33 | 19 | 14 | 17 | 18 |
| Quintile 2 | 20 | 27 | 23 | 18 | 16 | 16 |
| Quintile 3 | 20 | 19 | 24 | 22 | 18 | 18 |
| Quintile 4 | 20 | 13 | 20 | 24 | 22 | 21 |
| Quintile 5 | 20 | 7 | 15 | 23 | 28 | 27 |
| Region (%) |  |  |  |  |  |  |
| Northeast | 43 | 55 | 53 | 44 | 36 | 30 |
| Midwest | 15 | 10 | 17 | 17 | 13 | 19 |
| South | 18 | 14 | 15 | 20 | 22 | 17 |
| West | 12 | 11 | 5 | 8 | 16 | 20 |
| NDVI (in 270m distance buffer) | 0.38 (0.17) | 0.41 (0.19) | 0.39 (0.17) | 0.38 (0.16) | 0.37 (0.15) | 0.33 (0.15) |
| NDVI Quintile (%) |  |  |  |  |  |  |
| Quintile 1 | 18 | 17 | 15 | 15 | 18 | 29 |
| Quintile 2 | 19 | 15 | 17 | 20 | 22 | 22 |
| Quintile 3 | 19 | 15 | 19 | 22 | 22 | 20 |
| Quintile 4 | 19 | 18 | 22 | 22 | 21 | 16 |
| Quintile 5 | 20 | 32 | 23 | 19 | 16 | 11 |
| Neighborhood SES (time-varying) Quintile (%) |  |  |  |  |  |  |
| Quintile 1 | 20 | 33 | 21 | 15 | 15 | 15 |
| Quintile 2 | 20 | 20 | 20 | 20 | 20 | 20 |
| Quintile 3 | 24 | 23 | 24 | 25 | 24 | 23 |
| Quintile 4 | 12 | 10 | 11 | 12 | 13 | 15 |
| Quintile 5 | 24 | 14 | 24 | 28 | 29 | 27 |

<sup>a</sup> Value is not age-adjusted

**Table S4.** Hazard ratios (HR) and 95% confidence intervals (CI) for anthropogenic L_50_ nighttime and anthropogenic L_50_ daytime sound pressure (dBA) and cardiovascular disease incidence in the Nurses’ Health Study (n = 95,225; 2000-2018)

|  | L<sub>50</sub> nighttime noise |  | L<sub>50</sub> daytime noise |  |
| --- | --- | --- | --- | --- |
|  | Age and Calendar Year Adjusted HR (95% CI) | Fully Adjusted* HR (95% CI) | Age and Calendar Year Adjusted HR (95% CI) | Fully Adjusted* HR (95% CI) |
| **CVD (n = 6,448)** | 1.00 (0.97, 1.03) | 1.03 (1.00, 1.05) | 0.99 (0.97, 1.02) | 1.02 (0.99, 1.05) |
| **CHD (n = 3,108)** | 1.02 (0.98, 1.06) | 1.05 (1.01, 1.09) | 1.02 (0.98, 1.06) | 1.05 (1.00, 1.09) |
| **Stroke (n = 3,380)** | 0.98 (0.94, 1.01) | 1.01 (0.97, 1.04) | 0.97 (0.93, 1.01) | 1.00 (0.96, 1.04) |

\* Hazard ratios are adjusted for age and calendar year, race, smoking status, pack-years smoked, family history of MI, menopausal status, postmenopausal hormone use, diet, alcohol consumption, BMI, night shift work, parental occupation, educational attainment (RN degree), marital status, husband's highest education, nSES score quintile, region, and air pollution (PM<sub>2.5</sub>). Note: IQR for anthropogenic L<sub>50</sub> nighttime sound: 3.67 dBA; IQR for anthropogenic L<sub>50</sub> daytime sound: 4.35 dBA.

**Table S5.** Additional (for sensitivity analysis) Age-Adjusted Nurses’ Health Study Participant Characteristics by Quintiles of Nighttime Anthropogenic L_50_ dBA from 1988-2018 (n = 114,116)

| Characteristic | Total | Nighttime L<sub>50</sub> dBA Quintile 1 | Nighttime L<sub>50</sub> dBA Quintile 2 | Nighttime L<sub>50</sub> dBA Quintile 3 | Nighttime L<sub>50</sub> dBA Quintile 4 | Nighttime L<sub>50</sub> dBA Quintile 5 |
| --- | --- | --- | --- | --- | --- | --- |
| Comorbidities at baseline (diabetes, elevated cholesterol, and/or high blood pressure) (%) | 47 | 47 | 48 | 48 | 47 | 47 |
| Statin use (%) | 20 | 20 | 20 | 20 | 19 | 19 |
| Aspirin use, doses (%) |  |  |  |  |  |  |
| Non-user | 9 | 8 | 8 | 9 | 9 | 9 |
| Former user | 31 | 32 | 32 | 31 | 31 | 31 |
| Current user, 1-5 doses/week | 33 | 34 | 33 | 33 | 32 | 32 |
| Current user, 6+ doses/week | 10 | 10 | 10 | 10 | 10 | 10 |
| Missing | 17 | 15 | 16 | 17 | 18 | 20 |
| Physical activity categories (MET hours/week) |  |  |  |  |  |  |
| < 3 | 16 | 16 | 16 | 16 | 16 | 17 |
| 3 to <9 | 17 | 17 | 18 | 17 | 18 | 17 |
| 9 to <18 | 16 | 16 | 16 | 16 | 16 | 15 |
| ≥ 18 | 19 | 20 | 19 | 19 | 18 | 17 |
| Missing | 32 | 31 | 31 | 31 | 32 | 34 |

**Table S6.**
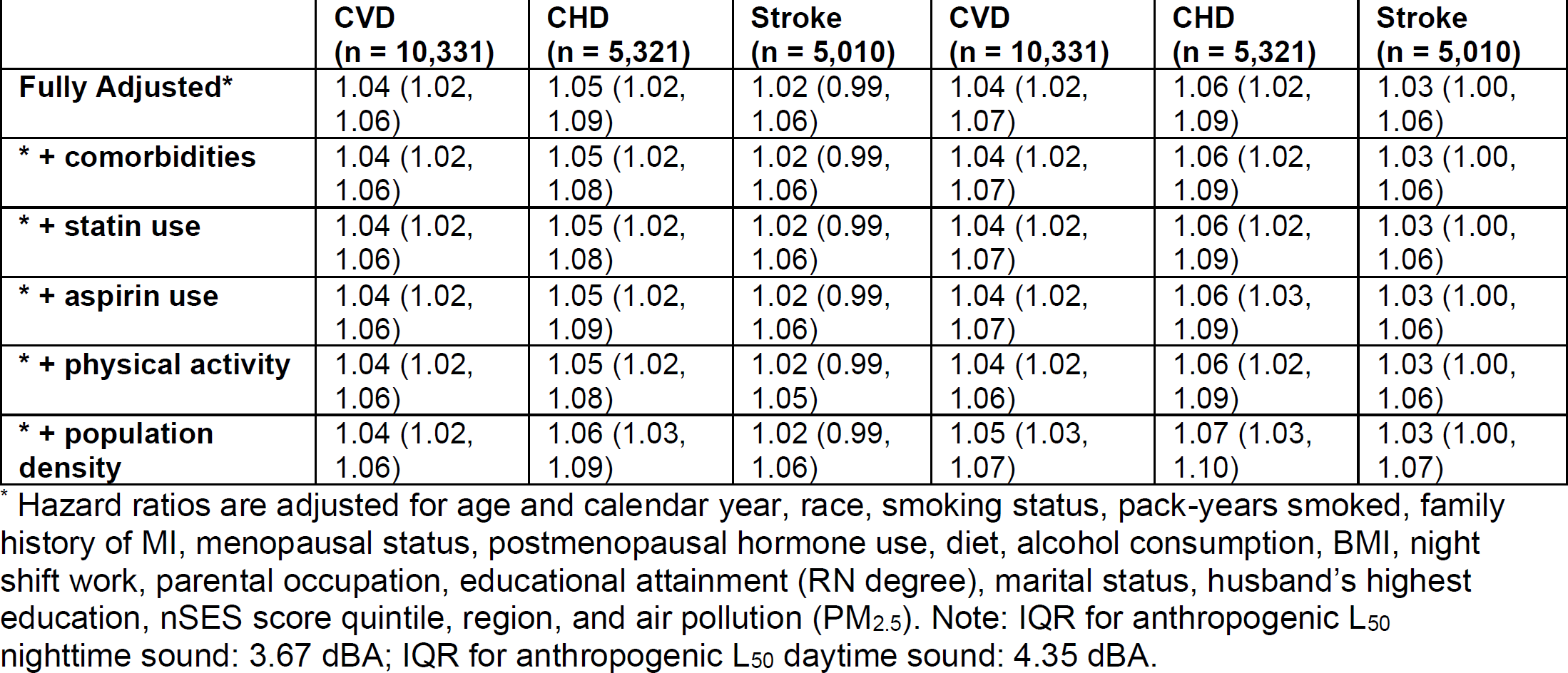
Hazard ratios (HR) and 95% confidence intervals (CI) for anthropogenic L_50_ nighttime and anthropogenic L_50_ daytime sound pressure (dBA) and cardiovascular disease incidence in the Nurses’ Health Study (n = 114,116; 1988-2018) with additional adjustment for comorbidities at baseline, statin use, aspirin use, physical activity, and population density.

**Table S7.** Estimated proportion of association of nighttime noise and CVD in the Nurses’ Health Study explained by short self-reported sleep duration.

| Mediator | Proportion of association of L<sub>50</sub> nighttime noise and CVD explained by mediator, with 95% confidence intervals | Proportion of association of L<sub>50</sub> nighttime noise and CHD explained by mediator, with 95% confidence intervals | Proportion of association of L<sub>50</sub> nighttime noise and stroke explained by mediator, with 95% confidence intervals |
| --- | --- | --- | --- |
| **Self-reported 5 or less hours sleep duration per night** | < 0.01% | < 0.01% | < 0.01% |

